# CAN A MACHINE LEARNING ALGORITHM IDENTIFY SARS-COV-2 VARIANTS BASED ON CONVENTIONAL rRT-PCR? PROOF OF CONCEPT

**DOI:** 10.1101/2021.11.12.21266286

**Authors:** Jorge Cabrera Alvargonzález, Ana Larrañaga Janeiro, Sonia Pérez Castro, Javier Martínez Torres, Lucía Martínez Lamas, Carlos Daviña Nuñez, Víctor Del Campo-Pérez, Silvia Suarez Luque, Benito Regueiro García, Jacobo Porteiro Fresco

## Abstract

Severe acute respiratory syndrome coronavirus 2 (SARS-CoV-2) has been and remains one of the major challenges humanity has faced thus far. Over the past few months, large amounts of information have been collected that are only now beginning to be assimilated. In the present work, the existence of residual information in the massive numbers of rRT-PCRs that tested positive out of the almost half a million tests that were performed during the pandemic is investigated. This residual information is believed to be highly related to a pattern in the number of cycles that are necessary to detect positive samples as such. Thus, a database of more than 20,000 positive samples was collected, and two supervised classification algorithms (a support vector machine and a neural network) were trained to temporally locate each sample based solely and exclusively on the number of cycles determined in the rRT-PCR of each individual. Finally, the results obtained from the classification show how the appearance of each wave is coincident with the surge of each of the variants present in the region of Galicia (Spain) during the development of the SARS-CoV-2 pandemic and clearly identified with the classification algorithm.

## 2 INTRODUCTION

There have been 235 million cases of and more than 4.5 million deaths associated with SARS-CoV-2, the causal agent of coronavirus disease 2019 (COVID-19) ^1^. The standard method for the detection of SARS-CoV-2 is based on real-time reverse transcriptase polymerase chain reaction (rRT-PCR) performed using a nasopharyngeal swab sample ^2^. Massive testing, in conjunction with other control measures, has been implemented to identify symptomatic or asymptomatic carriers to prevent the spread of SARS-CoV-2. The cycle threshold (Ct) value is inversely related to the amount of RNA of the virus present in the sample, which has awaked interest as an indirect method to predict infectivity, disease progression, severity and even associated mortality ^3^.

rRT-PCR tests are commonly considered qualitative tests (i.e., providing just a positive or negative result); however, they provide a Ct value for each target gene, which indicates the number of PCR cycles required to reach the threshold level of fluorescence associated with a positive result. Hence, the Ct value is inversely proportional to the viral load, although this correlation is not linear and depends on many factors. A recent work reviewed several works published on the connection of Ct with patient conditions and clinical outcomes^4^.

In addition, research has shown that although Ct is correlated with viral loads, it is not gene-independent. Mutations that affect the template sequence where the primers bind can affect the amplification efficiency and, therefore, the Ct values for a specific gene. For SARS-CoV-2, increases in Ct values or nondetection of different genes associated with specific mutations of the virus have been described ^5, 6, 7^. In fact, variant B.1.1.7 can be efficiently identified with specific tests through an undetectable S gene target or significant increase in the Ct value of the S gene compared to other targets^8, 9, 10^; nevertheless, a significant delay of the Ct to N gene was observed in another test ^11^, which confirms that the differences in Ct values between genes depend on the virus mutations but also on the primers used and, therefore, could be considered assay-dependent.

The mutation rates of coronavirus are low in general. However, in the case of SARS-CoV-2, the numbers reached in the pandemic have led to the accumulation of mutations and the emergence of multiple lineage and variants. Some of them are classified as variants of interest (VOIs), and variants of concern (VOCs)^12 13 14^, which have relevant epidemiological characteristics that may affect the virus’s properties, spreads, clinical characteristics of disease, and vaccine and drug performance. For this reason, it is important to track known variants and implement surveillance systems capable of detecting significant changes in the predominant variants, the variants causing an outbreak or even the emergence of a variant of high consequence (VOHC). The gold standard for identifying and tracking variants in circulation is the whole genome sequencing, although it has important limitations related to costs, resource availability, lack of expertise, standardization and data delay.

Currently, machine learning or deep learning models have been used in the medical field and, more specifically, in the field of COVID. In particular, several works have developed classification models based on X-ray images ^15 16^. Beyond image-based models, several works have addressed information from blood ^17^ or medical information ^18, 19^. More specifically, according to the present work, ^20^ present a model established to predict PCR results based on clinical information alone.

There are two main approaches: unsupervised learning and supervised learning. In unsupervised learning, we should allow the algorithm to seek its own way to classify the data; however, for the supervised approach, we must give the algorithm a target set of clusters. In this work, we opted for the latter.

The Microbiology Department of the University Hospital of Vigo (Complexo Hospitalario Universitario de Vigo, CHUVI), was a pioneer in the use of pooling in saliva for detecting SAR-CoV-2 in a nonsymptomatic population ^21^; and over the pandemic, the ‘pooling lab’ has screened more than 750,000 individual samples by pooling. When we analyzed the results of the positive samples in pools versus individuals (data not yet published), we observed how the relationship between Ct values remained constant for each sample despite the increase caused by dilution. These relationships between Cts seemed to be able to be grouped into similar profiles, and some groups seemed to have temporal accumulation. These findings and those explained in the previous paragraph made us consider whether there is an underlying ‘signature’ or ‘pattern’ in the Cts results of an rRT-PCR that can be used for the classification of samples.

The main goal of this work is to assess whether there is a Ct pattern that is characteristic of virus variants. By creating a sufficiently large database to efficiently train a classification algorithm, we demonstrate that there is an underlying signature response to the rRT-PCR of the main virus variants and that the Ct results of a standard rRT-PCR test can be efficiently employed to infer the most likely variant infecting an individual. Due to limitations in the number of tests completely sequenced, we will train our algorithm to predict the waves inside the evolution of the pandemic and then compare this prediction with the arrival and predominance of the different virus variants in the area where this study was conducted.

The study protocol 2021/295 was approved by the Galician network of committees of research ethics.

## 3 METHODOLOGY AND MATHEMATICAL BACKGROUND

### 3.1 Brief description of the rRT-PCR technique employed, primers, and software

We performed nucleic acid extraction in a MicrolabStarlet IVD platform using the STARMag 96×4 Universal Cartridge Kit (Seegene Inc, Seoul, South Korea). To detect SARS-CoV-2, we applied the Allplex™ SARS-CoV-2 Assay (Seegene Inc, Seoul, South Korea), a multiplex one-step rRT-PCR able to simultaneously detect four viral targets, including the structural protein envelope (E) gene, the RNA-dependent RNA polymerase (RdRP) gene, the spike (S) gene, the nucleocapsid (N) gene, and an exogenous RNA-based internal control (IC). This rRT-PCR step was run on a CFX96™ system (Bio–Rad Laboratories, Hercules, CA, USA), and the analysis was performed using Seegene Viewer-specific SARS-CoV-2 software (Seegene Inc, Seoul, South Korea), resulting in separate cycle threshold (Ct) values for the E and N genes and one combined Ct value for the RdRp and S genes (RdRp/S) in the FAM, Cal Red 610 and Quasar 670 channels, respectively. The HEX channel is used for internal control. Regarding interpretation of the results, according to the manufacturer’s instructions, Cts values ≤ 40 are considered detected, and Cts value >40 or not applicable (N/A) are considered not detected.

### 3.2 Description of the dataset employed in this work

Positive samples from two different sources were used for this study. First, 3,274 positive samples were obtained from the 688,763 samples processed by the pooling techniques between August 2020 and July 2021. Second, we also included 17,144 positive samples obtained from the 313,939 samples processed in the microbiology laboratory between February 2020 and March 2021.

The samples processed in the ‘pooling lab’ are screenings to detect SAR-CoV-2 in a nonsymptomatic population. The participants were asked to collect saliva (self-sampling) in TRANSPORT MEDIUM-2 (Vircell® Ref: TM013) immediately after waking up, following the manufacturer’s instructions. Although each result pertains to an individual rRT-PCR for each sample, these samples were first flagged as possibly positive by group testing. Then, the original samples from the positive pool are individually analyzed. These are the results that are used here. Individual samples and pools were analyzed following the same standard rRT-PCR protocol described in 3.1.

The other samples were nasopharyngeal swabs processed individually in the laboratory of the CHUVI Microbiology Departament as part of the assistance routine for SARS-CoV-2 diagnosis. It is important to note that the supply of this source of positive samples ended prematurely in March 2021 due to the need to change the reagent used in this laboratory (Allplex™ SARS-CoV-2 Assay to Allplex™ SARS-CoV-2/Flu A/Flu B/RSV assay, both from Segene Inc.) because of the high demand. In this way, we were able to keep the Allplex™ SARS-CoV-2 assay to group testing, since in this case, an assay change requires a full re-evaluation of the system, and the increase in the Cts for the N gene previously described by Wollschäger et. al.^11^ may have greater significance in group testing. As explained in Section 3.3, the data from 12,313 positive samples obtained by the Allplex™ SARS-CoV-2/Flu A/Flu B/RSV assay between February and August 2021 could not be included in the present study.

### 3.3 Characterization of the wave concept

Since the pandemic began in March 2020, the concurrent increases and decreases in cases have been linked to the concept of “wave, which are determined using subjective, unofficial criteria. To the best of the authors’ knowledge, this is an abstract nomenclature whose rigorous definition has not yet been clearly established to date. To characterize the pandemic dynamics in our area, we tracked the curve of active cases at the level of Galicia and, more specifically, Vigo and determined the boundaries between the so-called ‘waves’ in a data-driven way.

The database of active cases in the entire Galician region during the SARS-CoV-2 pandemic was obtained from data provided by the public health service of the Autonomous Spanish Community. In order to determine the time limits of each wave, the contagion curve is fit to a smoothed spline (R^2^=0.99), and the waves are defined by the local minima and maxima of the curve, as shown in Figure 1. Therefore, it can be concluded that although vaguely defined, waves are quite distinguishable, and the number of samples is inherently higher near the peak of each wave and much lower in their frontiers. Additionally, each new wave could also be associated with a higher proportion of samples with lower Ct values at the beginning ^22^.

**Figure 1:**
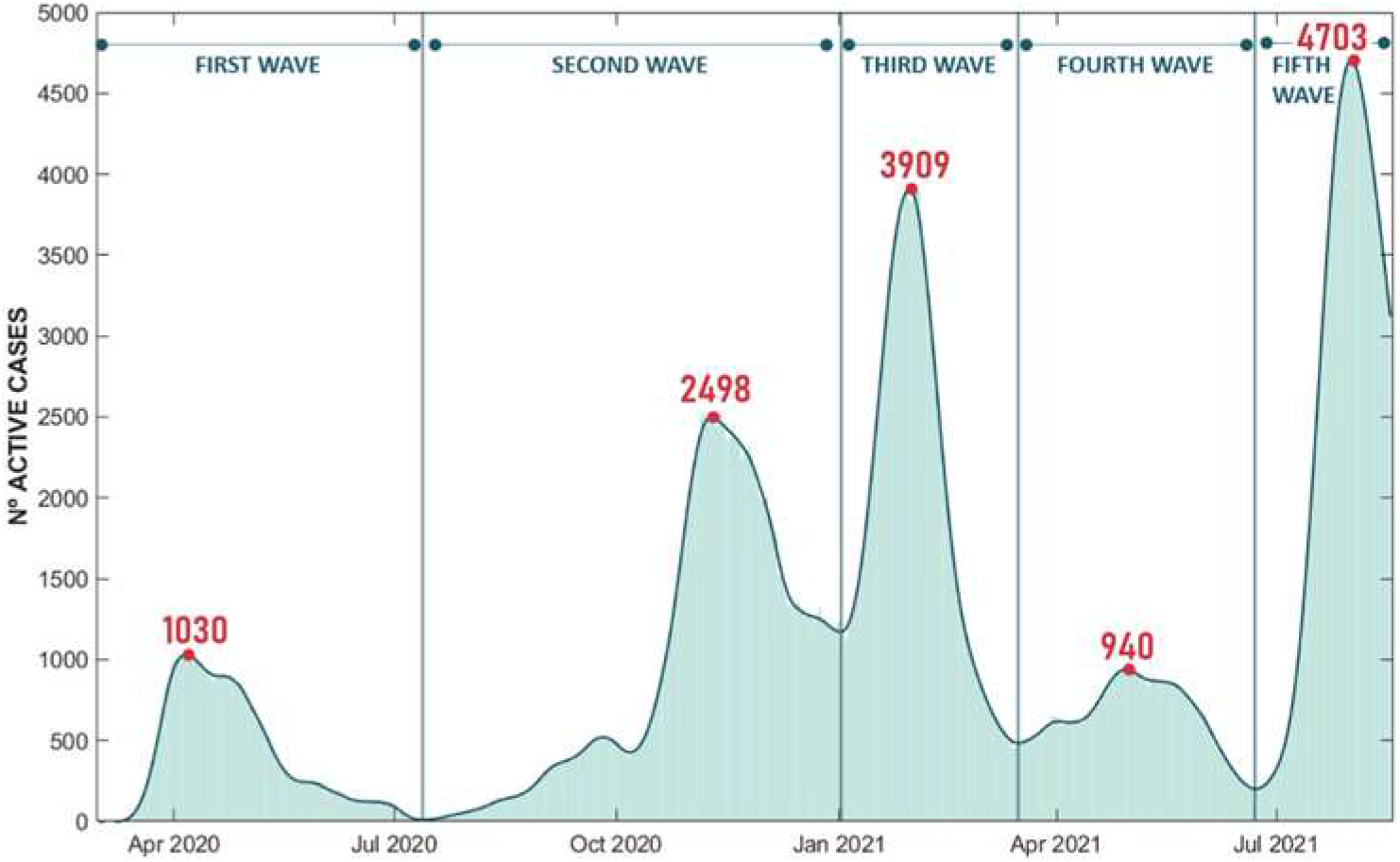
Smooth spline approximation of the infection curve in Vigo during the pandemic.

Focusing now on the data available for this study, **Figure 2** shows how the existence of the waves is reduced to four clearly differentiated peaks. First, the slight increase in the number of cases experienced in autumn of 2020 is not seen in the data collected. The peak of cases in the second wave is concentrated in the last months of the year.

**Figure 2:**
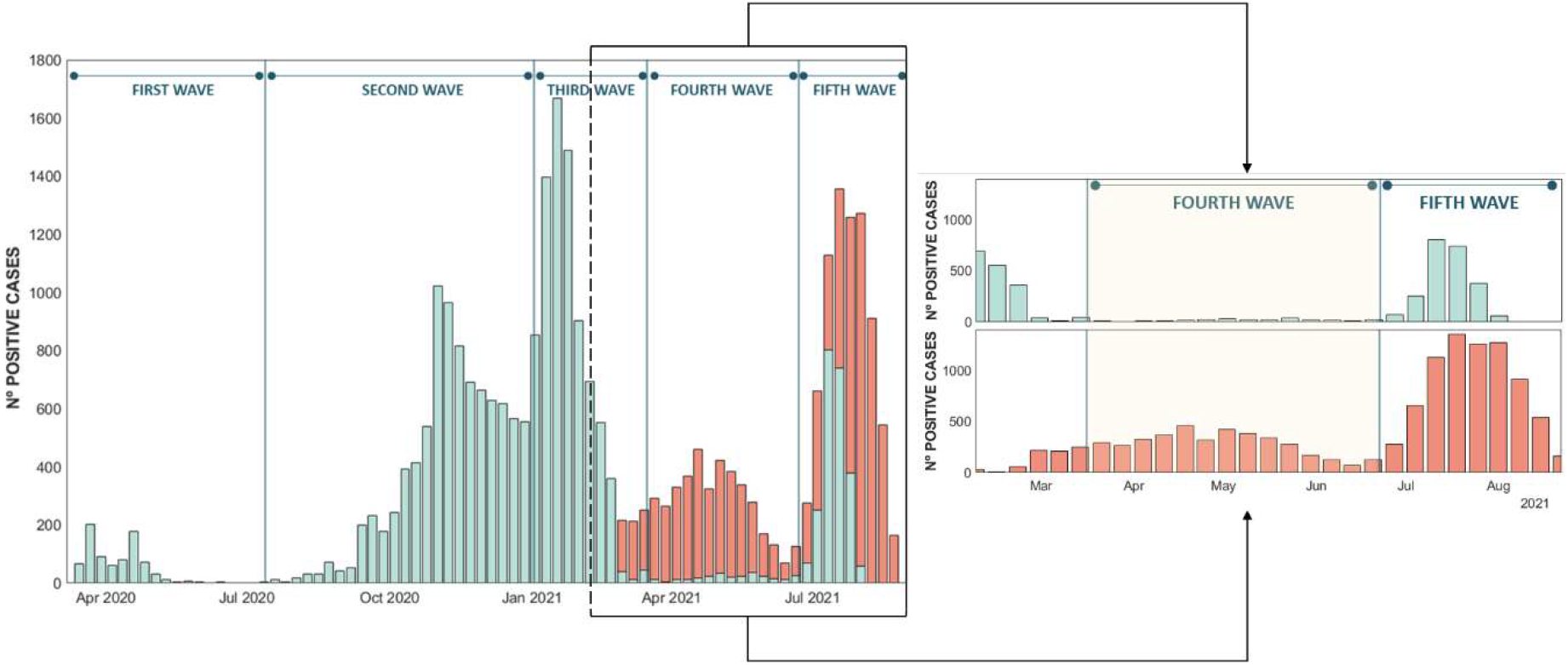
Number of positive SARS-CoV-2 tests in Vigo, averaged by week, detected by pooling in CHUVI, with the old PCR reagents (blue) and with the new PCR reagents (orange).

As will be explained in this work, the key aspect is the capacity of the machine learning algorithm to correctly predict the wave to which each sample belongs based on the numerical results of rRT-PCR for each gene. Therefore, a change in the target genes of the PCR performed during the fourth wave is too strong to indicate the temporal position of those tests and therefore had to be removed from our database to avoid giving an unfair advantage to the algorithm. Unluckily, the dire circumstances under which laboratories had to work during the pandemic led to this type of disturbance. Fortunately, in our case, it only significantly affected the fourth wave.

### 3.4 Descriptive analysis of the database used in the work

Even after extracting the samples that could lead to unfair results, the resulting database used in this study corresponds to a set of 20,418 PCR samples collected by the Microbiology Department of the CHUVI from March 2020 to July 2021. For each sample that tested positive for SARS-CoV-2 the database included an anonymized identification number, the date when the sample was taken, the threshold value for each target gene (E, N and RdRP/S) and the threshold value for the internal control (IC). The RdRP and S genes share the same channel; therefore, we obtained a single Ct value for both genes.

Figure 3. shows the distribution of the number of cycles from the analyzed gene profiles, where the average Ct value is approximately 26 for genes E and N and close to 28 for the combination of genes RDRP/S.

**Figure 3:**
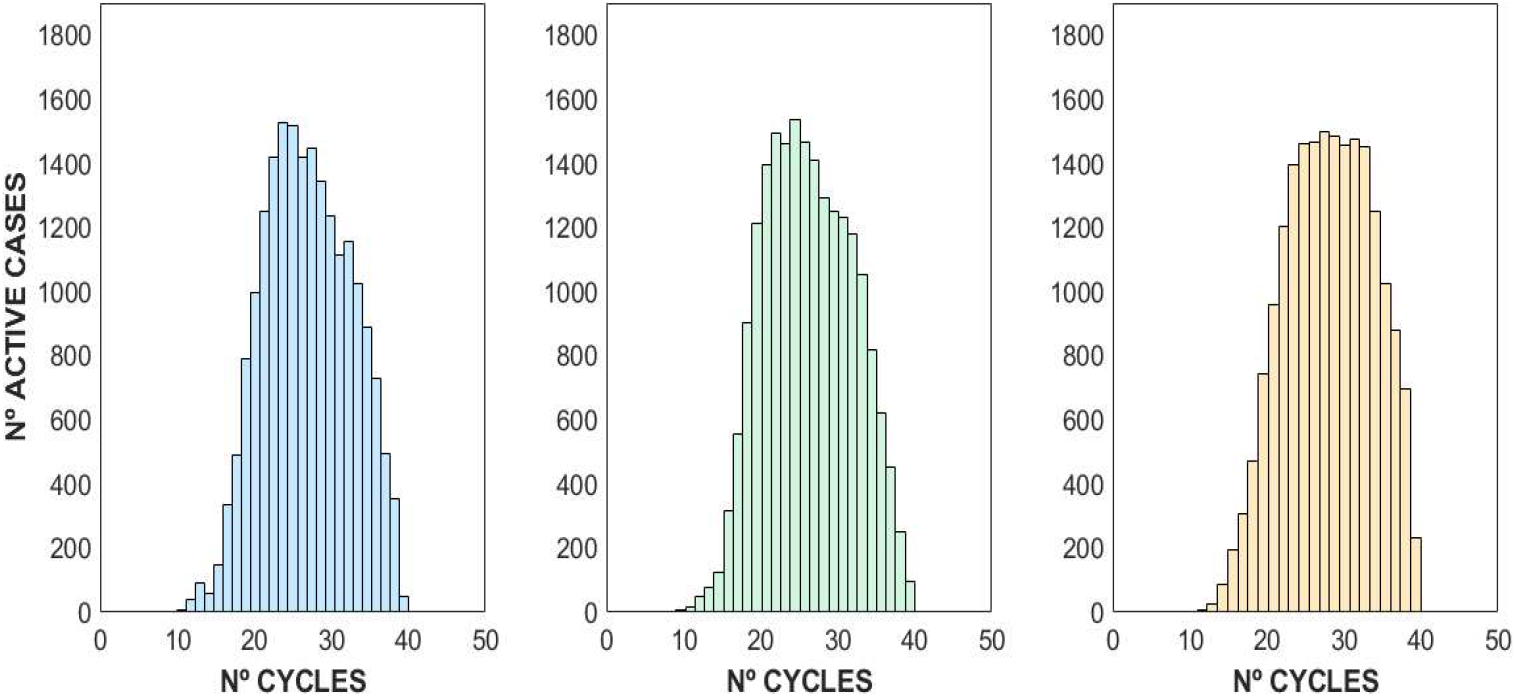
Gene E (blue), N (green) and RDRP/S (yellow) distributions from the pooling dataset.

Some visual features arise from the simple analysis of the data collected. As seen in **Figure 4**, the RdRP/S gene distribution seems to be slightly offset toward higher Ct values; and in fact, a more abrupt end is shown. However, a strong linear relationship between the number of cycles of the three genes can be observed from the database (R^2^_E-N_ = 0.96, R^2^_R-N_ = 0.95, and R^2^_E-R_ = 0.97). This is anticipated since the presence of the genes is expected to be similar and each number of cycles is allegedly related to the viral load of the individual; thus, the values of the numbers of cycles detected in any sample are usually quite close.

**Figure 4:**
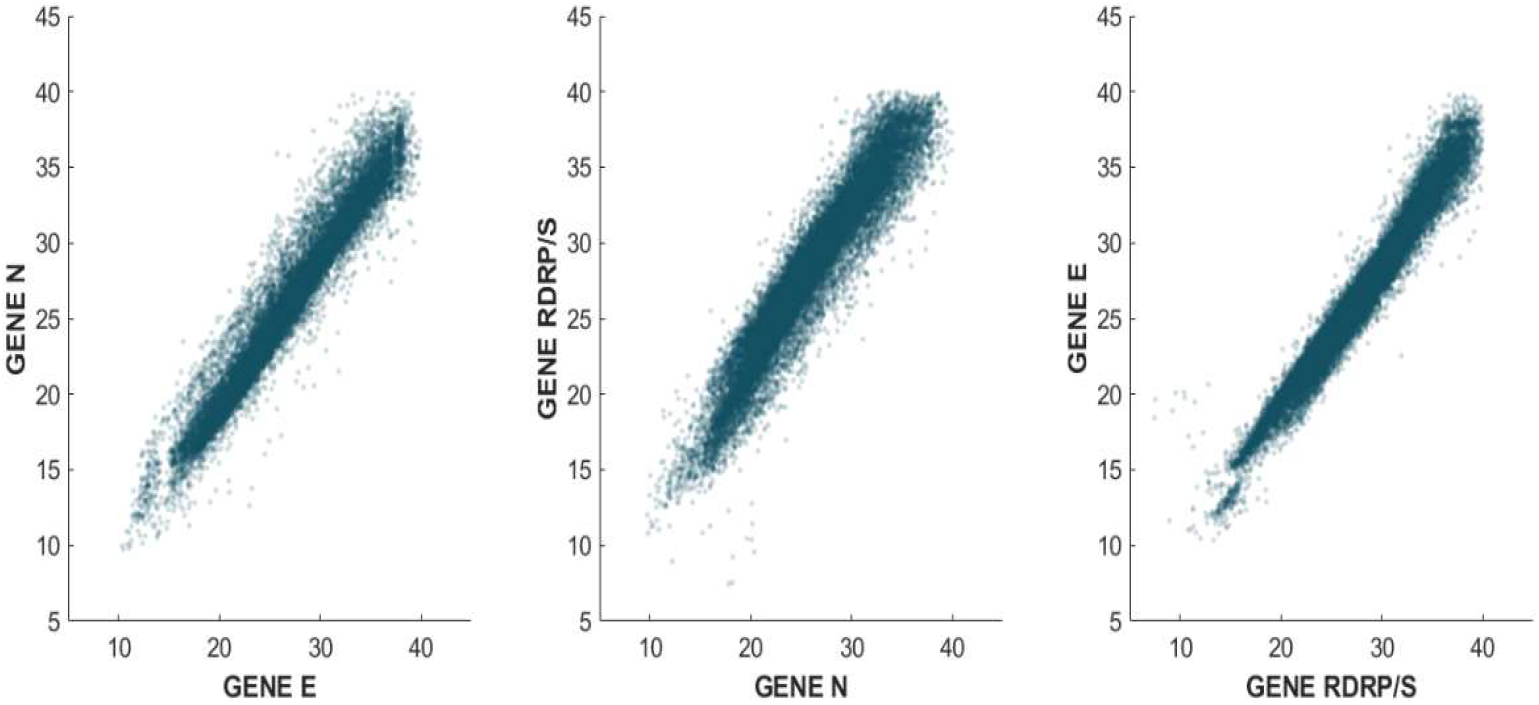
Relationship between the number of cycles of genes E, N and RDRP/S.

Figure 5. shows the temporal evolution of the number of cycles of each gene during the pandemic. The figure clearly shows that, at least at first glance, there is no trend or time evolution that points to Ct differentiation over time.

### 3.5 Working hypothesis

Since ML algorithms require a high volume of training data to be effective and genome sequencing for variant determination was impractical due to the high dimensionality of our database, we decided to use the wavenumber as the characteristic target cluster. Hence, the working hypothesis is the following: **each wave has a distinguishable pattern (signature) on the rRT-PCR results that allow an ML algorithm to efficiently predict the wave to which each individual test belongs**. If that hypothesis is proven true, some interesting conclusions can be extracted.

**Figure 5:**
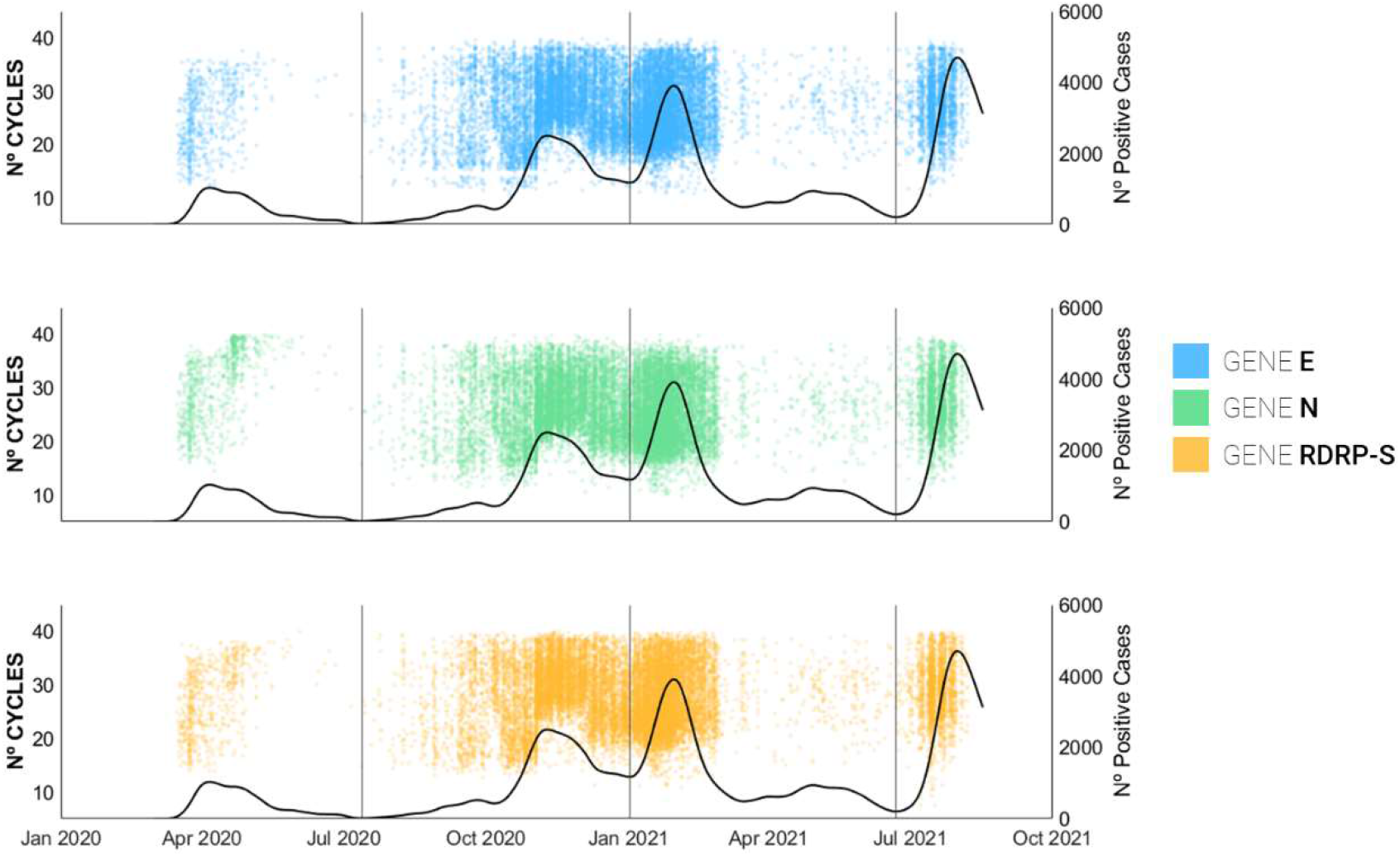
Evolution of the number of cycles of each gene against the number of positive cases during the pandemic.

**Figure 6:**
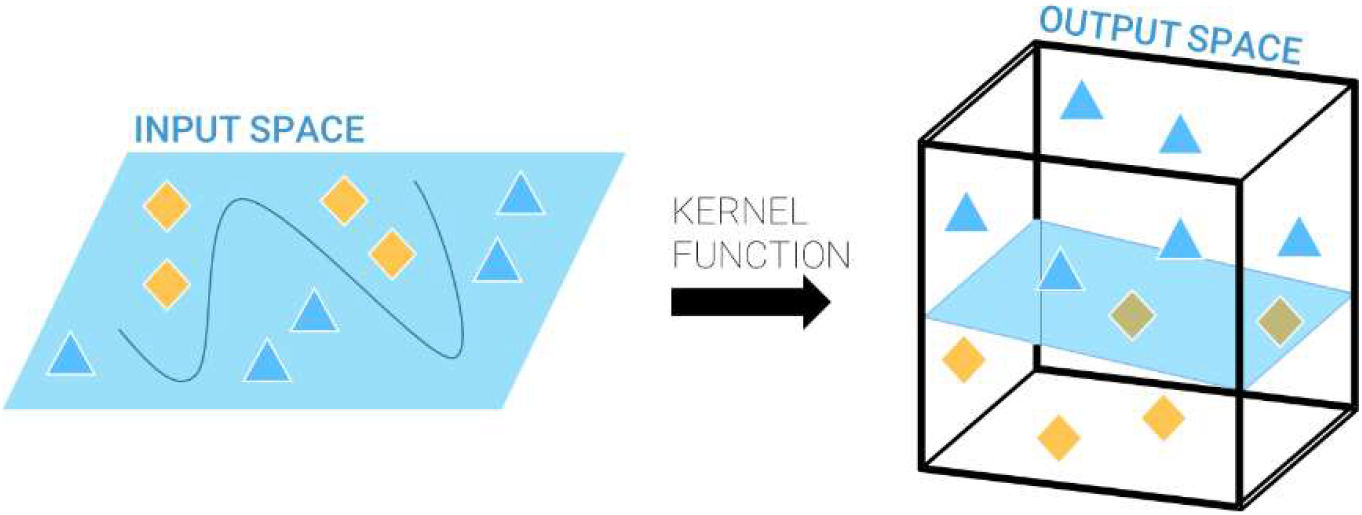
Support vector machine explanation.

**Figure 7:**
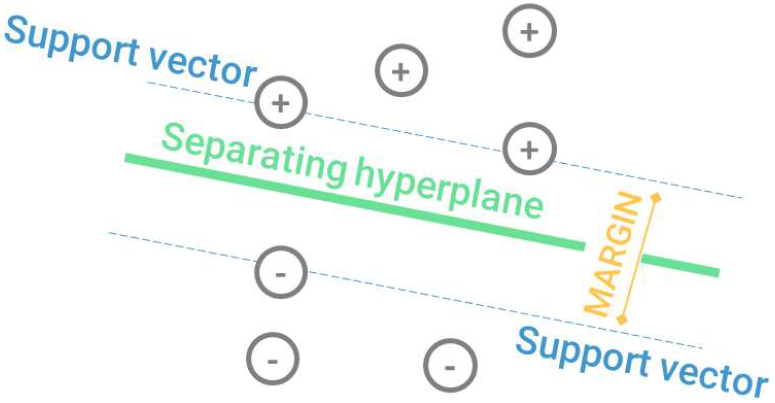
Maximum margin for a support vector machine.

To cluster the samples, a supervised learning technique would allow predicting the membership of a sample to a wave based simply on the number of cycles presented as a result of the PCR. Supervised learning algorithms are based on using labeled input data, i.e., with a correct answer with reference to its classification. Thus, as the algorithm is trained, it compares its predicted output with the correct input response until the error in its decision is minimized.

### 3.6 Classification techniques employed - SVM and NN

#### 3.6.1. Support Vector Machine (SVM)

From the diverse existing classification algorithms, the first option tested was a support vector machine (SVM), a kernel-based network that performs linear classification on vectors transformed to a higher dimensional space, i.e., it separates these vectors by means of an optimal hyperplane in the feature space that contains the main characteristics of the baseline data ^23^. According to the kernel trick ^24^, the creation of such a feature space is obtained by the transformation *ϕ* : *X* ⊂ *R*^*d*^ → *U* ⊂ *R*^*s*^, where *s* ≥ *d* (*d* represents the dimension of the original space, and *s* corresponds to the dimension of the feature space), allowing linear separating hyperplanes in the feature space to be equivalent to nonlinear separators in the original space.

SVMs tailored for classification are based on a sample *z*^*n*^, *z*_*i*_ =(*x*_*i*_, *y*_*i*_), where *x*_*i*_ *∈ X ⊂ R*^*d*^, *y*_*i*_ ∈ *Y* ={-1,1},∧*i*=1: *n* is linearly separable. I.e., it can be divided by a decision function *f*_*w,b*_ (*x*)=*sign* (*⟨ w, x ⟩* +*b*), *where w* ∈ *R*^*d*^∧*b* ∈ *R*. The optimal separating hyperplane is defined as the maximum margin separator hyperplane that maximizes its distance to the classes. It has two major weaknesses: the requirement of linear separability of the sample and their linear character.

In turn, this margin, or geometric margin, *τ* _*w, b*_, with respect to the sample *z*^*n*^ is described as

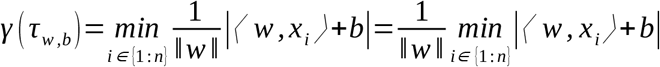

The optimal hyperplane is obtained as the solution of the following problem with its corresponding restrictions:

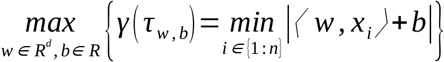

Considering that this is a multiclass problem since more than two input parameters are considered for training, it is important to differentiate two approaches: *one-vs*.*-all* and *one-vs*.*-one*.

- **<u>One-vs.-All</u>** ^25^: This approach is based on building *c* SVM models where the ith classifier is trained with all the examples of the ith class coded as +1 and the rest of the observations are coded ash −1 (hence its name one-against-all as each class is pitted against the rest).
- **<u>One-vs.-One:</u>** This methodology is based on the construction of *c(c - 1)/2* binary classifiers trained with elements of two classes 2 to 2. Thus, for each element of class i, the classification between the other elements of different classes can be established.

In the present work, both alternatives were tested. Since the SVM model used was optimized, it provided a better result in the one-versus-one application.

#### 3.6.2. Neural Networks

Neural networks were born from the field of biology in the search to create a system that would follow the learning patterns of the human brain. This was achieved through the work of psychiatrist McCulloch and mathematician Pitts ^26^, whose first model consisted of an input layer (containing the original data); an output layer (containing the classification result) and a certain number of hidden layers with their corresponding number of nodes connected by weights, which were assigned based on common characteristics. Mathematically, a neural network can be denoted by the function *f* : *X* ⊂ *R*^*d*^ *→Y* ⊂ *R*^*c*27^ that can be expressed as follows:

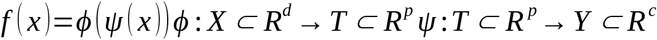

where *d* is the dimension of the input space, p is the number of neurons of the hidden layer, c is the dimension of the output layer, *T* is the hidden space, *ϕ* is the activation function of the hidden layer and *ψ* is the activation function of the input layer.

For the case of artificial neural networks called *multilayer perceptrons* (MLPs) that are characterized by having a series of neurons called *perceptrons*, a back propagation process that propagates the error back to the training in order to reduce the error until the NN learns the necessary information is used. Its expression can be shown as:

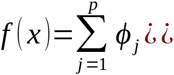

where *w*_*j*_ and *w*_0_ are the weights of the input layer and *c*_*j*_ and *c*_0_ are the weights of the hidden layer.

In this case, the neural network utilized was a feedforward, fully connected model specialized for classification (**Figure 8**). This means that all the neurons in each layer are connected to the neurons in the previous layer, with each fully connected layer multiplied by a weight matrix plus a bias vector that must be considered. Moreover, an activation function, which was previously mentioned, is placed between the layers, allowing nonlinear learning of patterns between layers. Finally, the last layer followed by the softmax activation function results in the network solution and consequent wave classification predictions.

**Figure 8:**
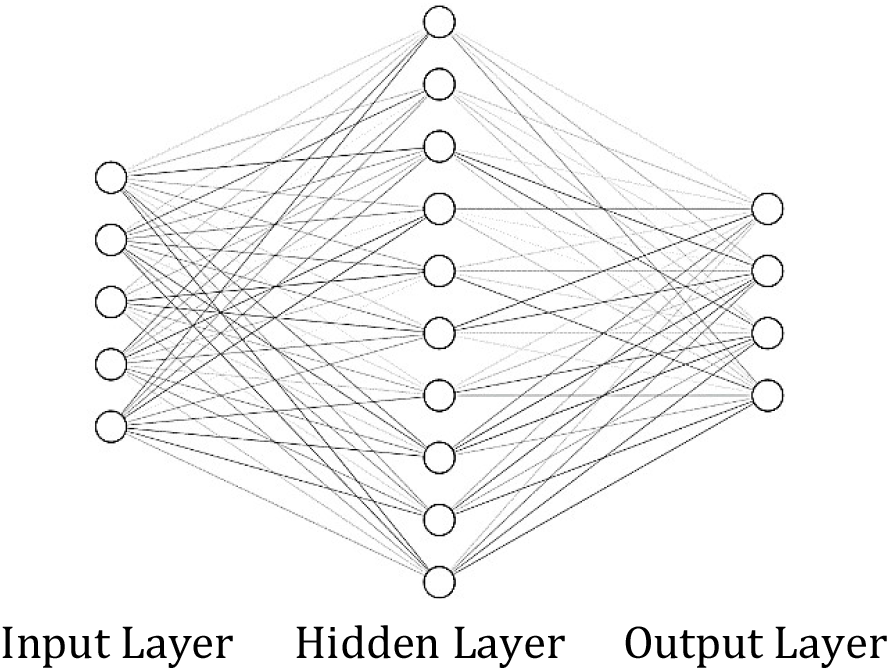
Feedforward, fully connected neural network structure.

#### 3.6.2. Structure of the model

Figure 9. includes the information detailing the several steps that constitute the entire ML pipeline. First, the number of cycles of each of the genes for a single sample and an additional number of cycles corresponding to the internal control are taken as input parameters. Then, the training process starts after the machine learning algorithm is chosen.

The output of the algorithm corresponds to a confidence score that represents the probability that a sample belongs to a particular group. Considering that the main objective is to predict the membership of a sample to a wave, the output of this algorithm will correspond to a confidence level associated with the probability that a sample belongs to a wave. Thus, the wave with the highest confidence level assigned to it will be the one chosen as the predicted wave.

**Figure 9:**
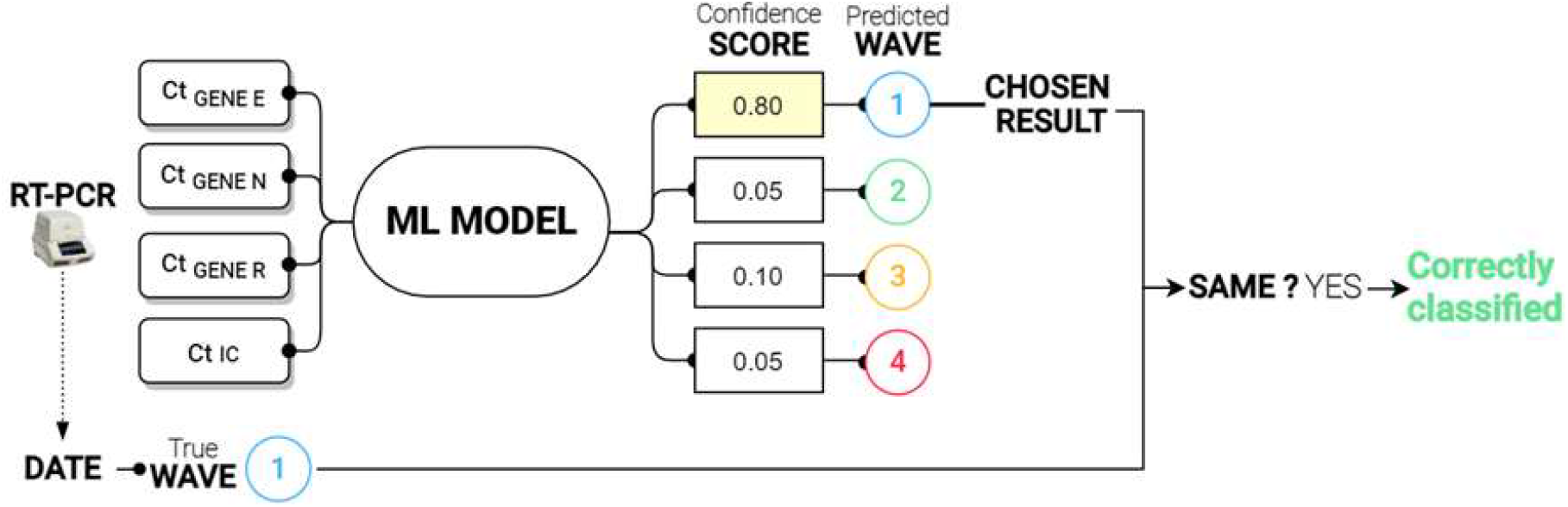
Predictive scenario of an ML model.

Subsequently, the prediction will be compared with the real wave value. If the prediction coincides with the real value, it represents a good estimating; conversely, it represents an incorrect classification. The actual wave value of the sample is determined from the date of the sample taken and the estimated cutoffs with the approximation of the wave of active cases to a spline.

## 4 RESULTS

This section will discuss the results obtained during the present investigation. The section will begin with the characteristics of the algorithms used, then address choosing the best alternative by evaluating the outcome and end with some conclusions drawn from the analysis.

### 4.1 Metrics

First, for the training of both models, a technique called *cross-validation* was utilized to avoid overfitting (i.e., the situation in which the network overlearns and extracts some noise as the main structure of the data, which affects the generalization of the predictions that tend to be incorrect). The idea behind cross-validation is to divide the database into a number of randomly chosen partitions, usually balanced, in order to train the model on subsets that it has not seen before. Thus, in this case, the database was divided into five parts. Four parts were used for training and the remaining part was used for testing. Then, the part used for validation was alternated as each training step was completed.

The criteria used to compare the results of the supervised algorithms are two widely known basic metrics: the *accuracy* and the *mean absolute error* (i.e., MAE). Considering that cross-validation was utilized, the accuracy was calculated as the percentage of observations correctly classified considering only the number of samples held out for validation in each training segment. Moreover, the MAE measures the average magnitude of the absolute differences between the prediction and the real value using Equation 4.1:

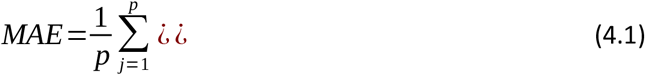

where *p* represents the total number of incorrectly classified samples, *y* _*j*_ is the prediction from the algorithm and 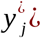 is the real value. As a linear score, all individual differences are weighted equally in the average, and the result include the units of the variable the model is seeking to predict. For this case, a difference of one unit corresponds to a prediction correlated to the next or previous wave with respect to the correct wave. Therefore, if the MAE falls close to unity, it will be interpreted as the algorithm having the tendency to mostly fail between contiguous waves where pattern differentiation is more complex.

Among the diverse metrics used to evaluate a model, the confusion matrix is often used to evaluate the results of a model and identify its “weak points”. In this matrix, the diagonal shows the percentage of correctly identified results (i.e., second waves identified as such), and the off-diagonal elements show the failure rates.

Furthermore, the confusion matrix also shows the *true positive rate* (i.e., the TPR) and *false negative rate* (i.e., the FNR) in the right-hand columns. The TPR is the proportion of samples correctly classified with respect to their true class, and the FNR is the proportion of samples incorrectly classified with respect to their true class.

### 4.2 Overview of the performances of the SVM and NN

The classifications made with both algorithms obtained similar results. This can be seen in the accuracy results in ***Table 1***. This similarity was expected from the linearity observed in the data, which led to equivalent results for both approaches. The cases in which the SVM shows a real improvement over the other techniques are usually related to nonlinear trends among the original data, which was not found in the present study.

**Table 1:** Classification algorithm specifications and training results.

| SUPPORT VECTOR MACHINE |  | NEURAL NETWORK |  |
| --- | --- | --- | --- |
| MODEL CHARACTERISTICS |  | MODEL CHARACTERISTICS |  |
| Kernel function | Gaussian | N° Fully Con. Layers | 1 |
| Kernel scale | 2.22 | First layer size | 100 |
| Box constraint level | 3.89 | Activation | ReLU |
| Multiclass method | One-vs-One | Regularization ($\lambda$) | 0 |
| TRAINING RESULTS |  | TRAINING RESULTS |  |
| Accuracy | 78.50 % | Accuracy | 78.60 % |
| MAE | 1.07 | MAE | 1.08 |

***Table 1*** includes the model characteristics for the *SVM*, following a one-vs.-one multiclass method with no standardization of the data; and for the *NN*, which has a unique hidden layer with 100 neurons and uses the ReLU activation function. It is important to note that despite the large difference in training times between the two algorithms, their accuracies are practically identical (this can be checked by examining the accuracies and MAEs in the same table).

***Figure 10*** and ***Figure 11*** include the confusion matrices for the algorithms, where the blue color is related to responses that are correct and the orange tone is related to misclassified samples. The fourth wave is the most clearly identified, with an accuracy of over 94% in both cases. However, the time frame between the second and third waves, as well as the transition from the first to the second, causes more confusion in the algorithms. As seen in the TPR column, the lowest hit percentage corresponds to the first wave in the case of the SVM (***Figure 10***) and to the third wave in the case of the NN (***Figure 11***).

**Figure 10:**
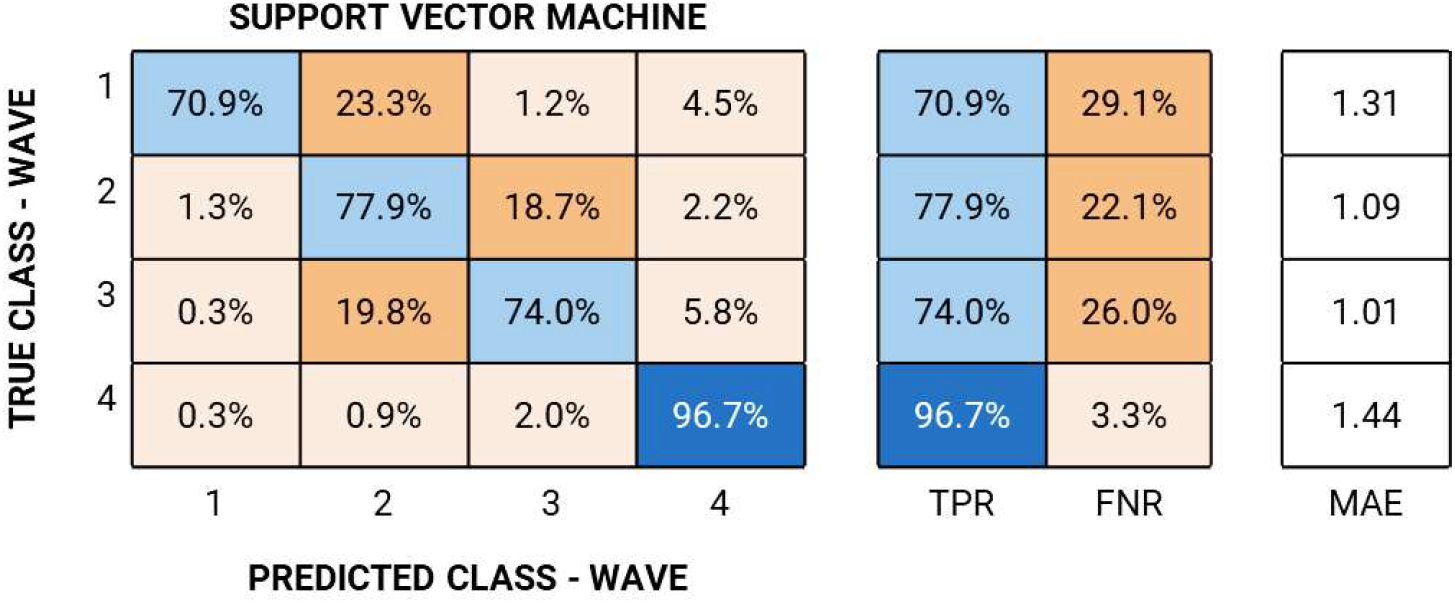
Support vector machine results.

**Figure 11:**
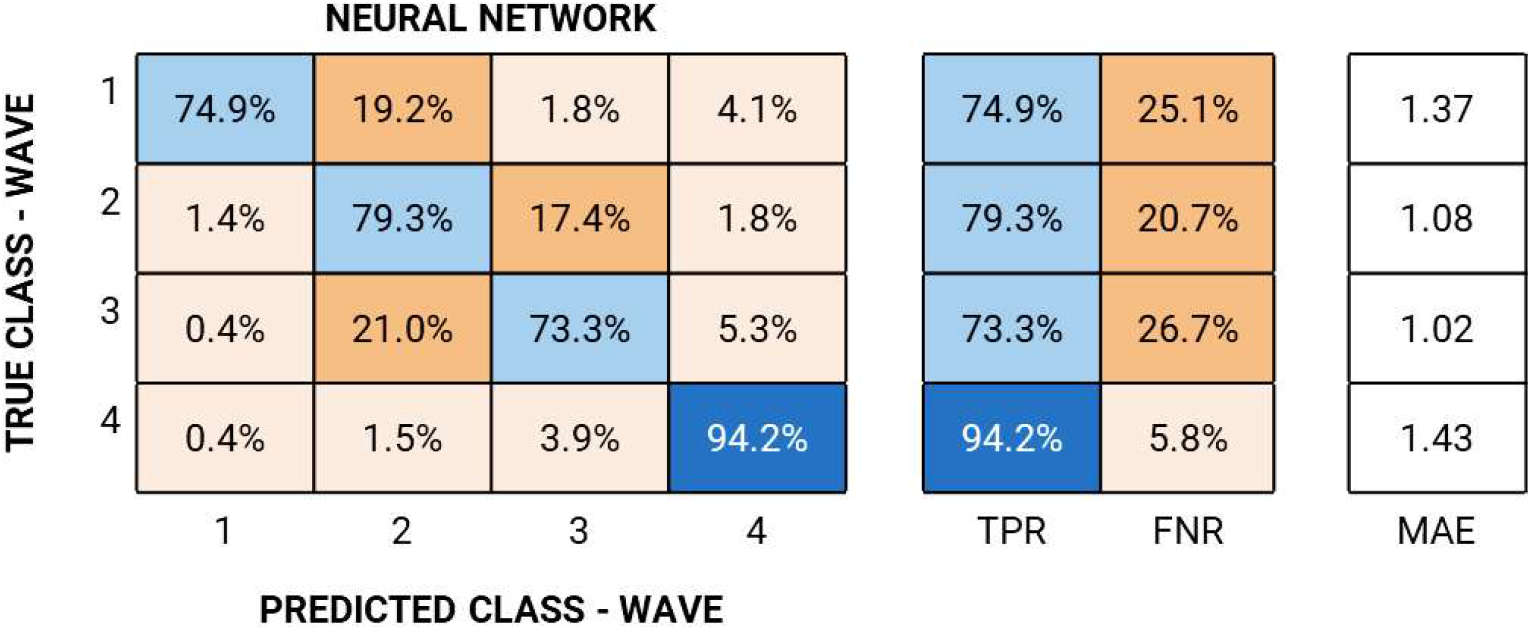
Neural network results.

Focusing now on the MAEs, it is worth highlighting the fact that the fourth wave, with the highest accuracy, is the one with the highest MAE, which means that those points that have been misclassified are usually associated with the first or second wave rather than the contiguous wave.

### 4.3 Classification results

When discussing the results, the outcome offered by the SVM is considered since they showed quite similar results and the SVM is a model that has certain advantages over the neural network in terms of performance and extrapolation of results. One such advantage is the opportunity to avoid retraining the model with the input of new samples, which is required in the case of the neural network.

Figure 12 shows the result of the classification by the SVM divided by waves, highlighting the predicted wave class assigned by the algorithm to the sample using color. The continuous black line shows the evolution of the number of infections throughout the pandemic (i.e., the number of positive cases) and the scattered points in time show the average confidence with which the classifier made the decision each day. That is, the average confidence is a ratio that shows the confidence with which the classifier assigns a wave to each individual.

**Figure 12:**
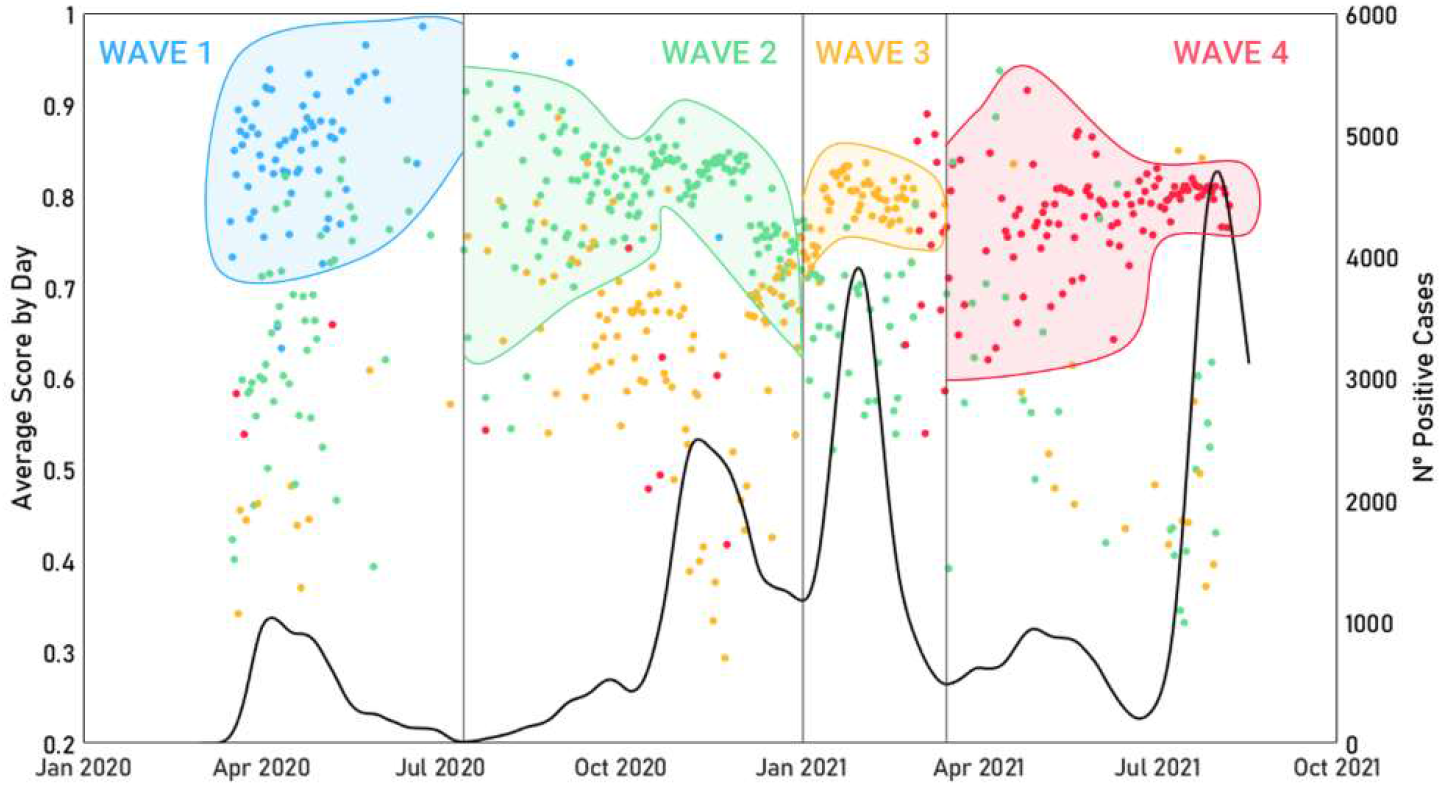
SVM classification result for the data utilized.

As seen in Figure 12, the majority of individuals are classified within their wave, and those points (located in the middle-bottom area of the figure) that have been misclassified are usually assigned to the upcoming or preceding wave.

An example of this confusion can be seen clearly between the second and third waves, where the yellow dots (individuals from the second wave that have been identified as third) can be identified within the second wave region. This can also be noticed, to a lesser extent, in the section of the first wave where a small cloud of green dots is located in the lower area (that is, real first one individual wrongly predicted by the classifier as second wave samples).

Moreover, Figure 13 shows how the increase in failures is related to a higher uncertainty in the solution obtained by the classifier. It should be considered that since the classifier has four possible answers, if a decision is made with a score close to 0.25, it means that all four options are necessarily similar in terms of score level. In contrast, those answers obtaining greater certainty (i.e., with a confidence level close to unity), correspond to a higher number of correct classifications and almost no confusion. Hence, the algorithm seems to be well aware of its real performance.

**Figure 13:**
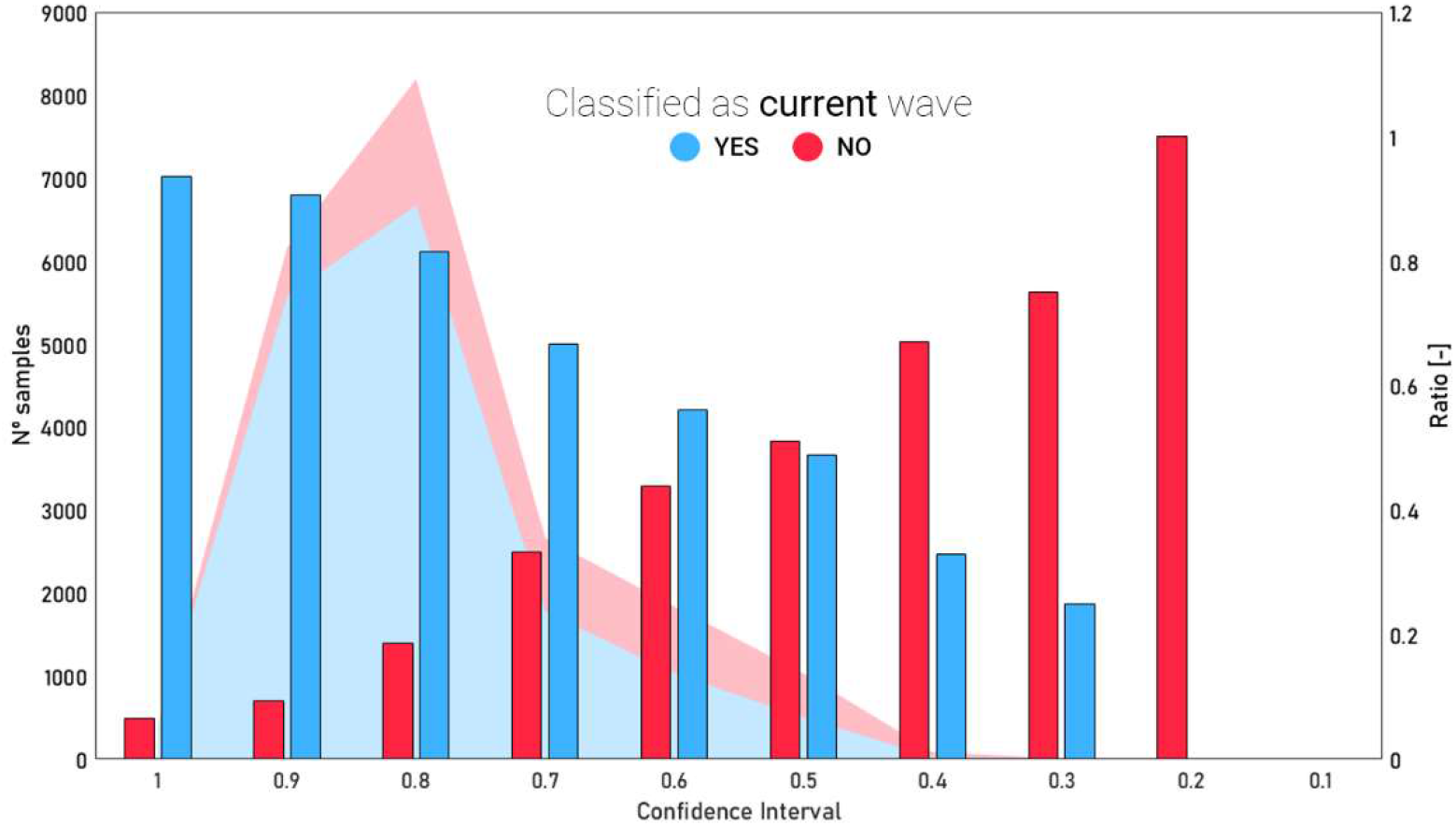
Ratio as a function of the confidence intervals for the SVM results.

It is worth mentioning that since the ground truth for each sample is based only on the date of their tests, which in the end is converted into a class membership (wave), the overlapping waves and the underlying causes that will be discussed later can easily be explained the fact that some (maybe many) of the individuals misclassified by ML could be in fact individuals misclassified to their assigned waves.

In fact, the algorithm precision tends to be higher at wave centers where the sample characterization is more solid. In contrast, those points that are located near the frontiers of the waves tend to be more conflicting (which can be easily seen, for example, in the intersection between the second and third waves in Figure 13).

## 5 DISCUSSION

Based on the results obtained in the classification, a deep reason to justify how a mathematical model, using only the detected number of cycles of each gene together with the number of cycles detected by the internal control as input data, could have such a high and accurate precision rate for each wave (as shown in the gray areas of Figure 14) was sought.

**Figure 14:**
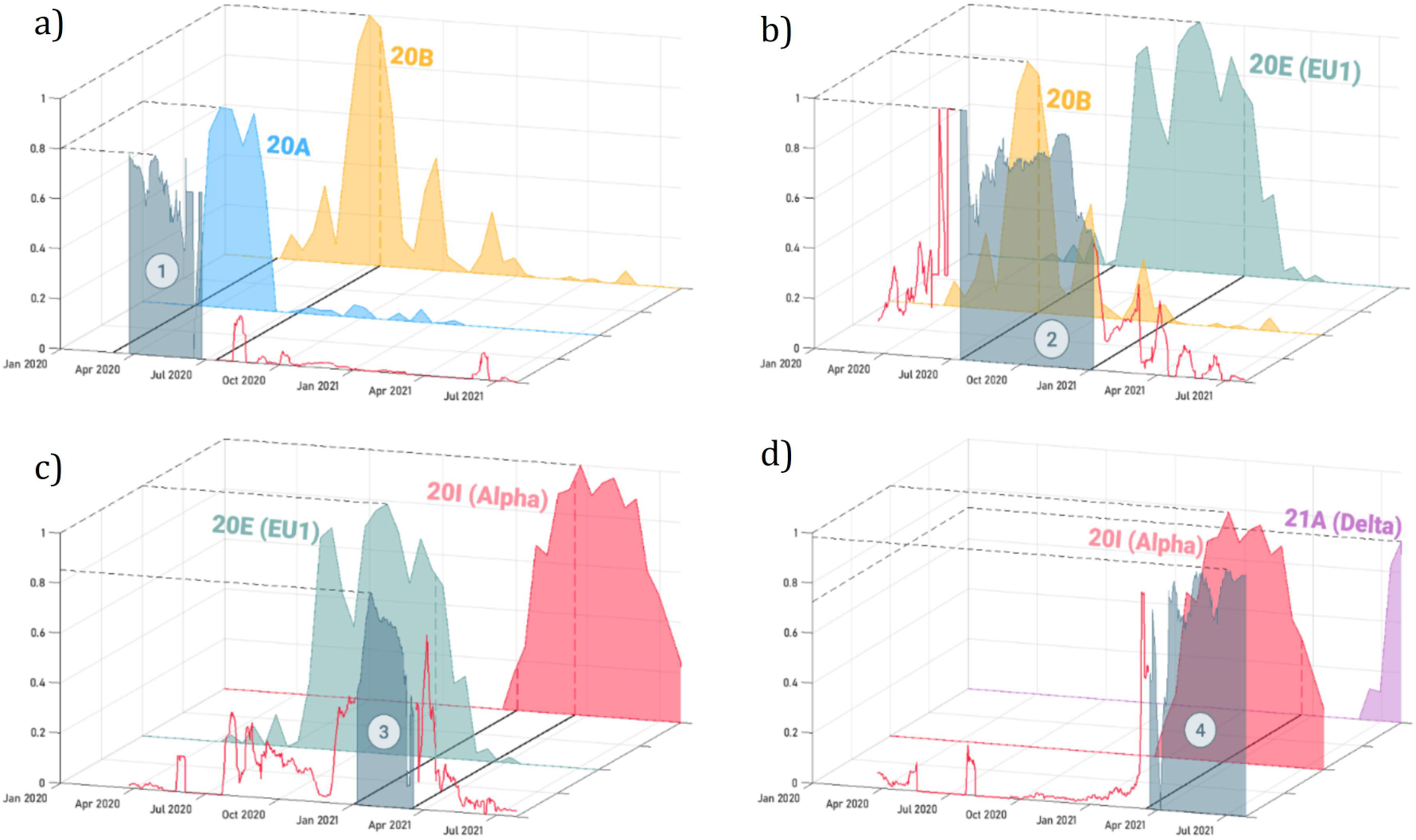
Accuracy compared to the prevalence of variants throughout the pandemic divided into the four waves (a, b, c and d, respectively), where the gray area is identified as the rate of correctly classified samples, and the solid red line represents the rate of incorrect classifications.

Starting with the results corresponding to the first wave (Figure 14 a)), the accuracy area (whose maximum reaches 80% at the peak of the wave) coincides temporally with the appearance of variant 20A in the Galician region. More specifically, the end of the wave also coincides temporally with the peak and decrease in active variant 20B cases (dashed line in the yellow area). This indicates that in addition to being characterized by variant 20A, this wave also picks up some features of variant 20B that cause a spike in the failure rate in October 2020 and again in July 2021.

In the case of the second wave (Figure 14 b), the tendency of the accuracy rate to follow the presence of specific variants still persists with the appearance of 20E (EU1) at the start of the second wave. Even so, this wave shows a higher error rate prior to its beginning and once it has ended since the 20B variant is present simultaneously with 20E (EU1) throughout the entire wave. Again, the figures show how the peaks in the error rate (red solid line) correspond approximately with spikes of 20B. Furthermore, the figures also show that from January 2021, the error rate decreases as the 20E (EU1) variant disappears.

The third wave (Figure 14 c), once again, its characterized by the presence of the 20E(EU1) variant together with 20I. This again causes a certain spike in the error rate due to the presence of these same variants during the rest of the waves. However, it is clearly observed that a higher accuracy corresponds to the coincidence of a higher percentage of cases of these variants simultaneously.

Finally, the fourth wave (Figure 14 d) is more clearly identified than the rest because the 21A variant is only present during the summer months of 2021. This means that the failure rate remains practically null until the arrival of this wave in April 2021.

### 5.1 Limitations

The main limitation of this study is that it is applied only to the data from a single area. This is justified by the fact that the dataset required for training had to avoid any trace of data that could lead the algorithm to distinguish data from other data. In fact, as mentioned above, this reduced the size of our dataset since we had to discard many tests due to the use of a different set of target genes. The purpose of this work is to show that a distinguishable signature on the Ct pattern seems to exist, but until proven using different labs, test conditions, etc., no further generalization should be made than the mere existence of this pattern.

### 5.2 Assessment of the potential interest of the proven concept

The ML tool proposed in this work represents an additional tool that can improve the relevance of rRT-PCR results. The classical interpretation of a qualitative, Boolean result (positive or negative) can be completed with additional information regarding the estimation of probable virus variants (if recognized by an ML algorithm). This can be useful for many purposes, such as the following: pandemic control (quick detection of the arrival of new variants), as a screening tool for virus sequencing, as a quality check of the tests and/or reagents, etc.

In addition, this work is just the initial step toward a completely new methodology applied to rRT-PCR not only in the case of SARS-CoV-2 but also in many other diseases.

## 6 CONCLUSIONS

In this work, we found that an ML algorithm trained with a sufficiently rich database can efficiently identify the moment in the SARS-CoV-2 pandemic when an individual was infected based on a simple, standard, rRT-PCR test with three channels (E, N and RdRP/S). No additional information regarding gender, age, condition, etc. was required by the algorithm. The subjacent reason for the precision of the ML algorithm seems to be an underlying characteristic signature of the main SARS-CoV-2 variants. As shown, the prediction seems to be aligned with the arrival and retreat of the different variants from the region where the tests were performed. Only by collecting a sufficient amount of data of different variants, individuals, tests, laboratories, etc. can the concept presented here be proven directly and not through the wave clustering concept employed here.

The results of this work can be a first step toward a new accessible and inexpensive surveillance method for tracking and/or selecting candidate samples for sequencing. Even with its limitations, this method may help monitor the changes to the virus and extend the surveillance to areas where current systems are scarcely implemented and have contributed significantly to the expansion of VOCs.

## Data Availability

All data produced in the present study are available upon reasonable request to the authors

## Notes

### Competing Interest Statement

The authors have declared no competing interest.

### Funding Statement

This study did not receive any funding

### Author Declarations

The Ethics committee/IRB os Servicio Galego de Saude gave a ethical approval for this work

